# Low rate of daily smokers in patients with symptomatic COVID-19

**DOI:** 10.1101/2020.06.10.20127514

**Authors:** Makoto Miyara, Florence Tubach, Valérie Pourcher, Capucine Morélot-Panzini, Julie Pernet, Julien Haroche, Said Lebbah, Elise Morawiec, Guy Gorochov, Eric Caumes, Pierre Hausfater, Alain Combes, Thomas Similowski, Zahir Amoura

**Author notes:** To whom correspondence shoul dbe sent: Pr Zahir AMOURA Alternative proof reader: Dr Makoto MIYARA and Pr Florence TUBACH. Contributed equally (Makoto Miyara, Florence Tubach).

## Abstract

**Background:** Identification of prognostic factors in COVID-19 remains a global challenge. The role of smoking is still controversial.

**Objective:** To evaluate the rate of daily smokers in patients with COVID-19.

**Methods:** COVID-19 in-and outpatients from a large French university hospital were systematically interviewed for their smoking status, use of e-cigarette and nicotinic substitutes. The rates of daily smokers in in-and outpatients were compared to those in the 2019 French general population, after standardization for sex and age.

**Results:** The inpatient group was composed of 340 patients, median age 66 years: 203 men (59.7%) and 137 women (40.3%), median age for both 66 years, with a daily smokers rate of 4.1 % CI95% [2.3–6.9] (5.4% of men, 2.2% of women). The outpatient group was composed of 139 patients, median age 44 years: 62 men (44.6%, median age 43 years), and 77 women (55.4%, median age 44 years). The daily smoker rate was 6.1 % CI 95% [2.7 - 11.6] (5.1% of men, 6.8% of women). In the 2019 French population, the daily smoker rate was 24.0% (27.5% of men, 20.7% of women). Among inpatients, daily smokers represented 2.2% and 3.4% of the 45 dead patients and of the 29 patients transferred to ICU, respectively.

The rate of daily smokers was significantly lower in COVID-19 patients, as compared to that in the French general population after standardization by age and sex, with Standardized Incidence Ratios of 0.24 [0.12-0.48] for outpatients and 0.24 [0.14-0.40] for inpatients.

**Conclusion:** Daily smokers rate in patients with symptomatic COVID-19 is lower as compared to the general population

## Introduction

As the pandemic of COVID-19, caused by the severe acute respiratory syndrome coronavirus 2 (SARS-CoV-2), is still under progression, the identification of risk factors is a global challenge. Among epidemiological risk factors, the role of smoking, to date, is unclear. Smoking has been initially found associated with adverse disease prognosis of COVID-19[1], although this finding remains controversial[2]. Reported rates of current smokers among SARS-CoV-2-infected patients range from 1.4% to 12.5% in China[1, 3-10], from 1.3% to 5.1% in the USA[11, 12], mainly for hospitalized patients (see systematic review in[13]). For outpatients, data are very scarce but also suggest similar low rates[13]. At first approach, the rates of current smokers in both COVID-19 in- and out patients seem to be low compared to the general population. These data notwithstanding, no firm conclusions can be drawn from these available COVID-19 studies because main potential confounders for smoking rate, namely age and sex, were not taken into account. Additionally, these studies included mostly hospitalized patients, and the low rate of current smokers may be related to high rate of patients with comorbidities (smokers having been advised to quit). Furthermore, these studies used data collected in the context of care in the medical files, which favors underreporting (patients being considered as non smokers when smoking status is not reported in the medical file) particularly when data collection is made by overwhelmed care healthcare teams for a disease *a priori* not related to smoking, and biased reporting (preferential smoking status collection in patients with pulmonary or cardiovascular comorbidities).

Therefore, the effect of current smoking on the risk of SARS-CoV-2 infection has yet to be determined. To accurately evaluate whether or not current smoking is associated with the risk of COVID-19, we conducted an observational study specifically designed to investigate this association, and compared the rates of daily current smokers after standardization by sex and age of two COVID-19 patients’ groups, one composed of outpatients (not subsequently hospitalized) and one of hospitalized patients (inpatients), with those reported in the 2019 French general population[14]

## Material and methods

### Patients and design

This is a cross-sectional survey specifically designed to investigate the smoking status of patients with COVID-19, both in hospitalized patients (representing the severe symptomatic cases of COVID-19) and in outpatients (i.e. patients who represent the non-severe symptomatic cases of this infection). Daily current smoker rates were compared to those of the 2019 French population as a reference, after standardization by age and sex.

Eligible patients were those with a confirmed diagnosis of COVID-19 at the APHP Pitié-Salpêtrière Hospital, France, with two groups: the inpatients: those hospitalized in medical wards of medicine (not including ICUs, as most patients cannot be adequately interviewed), and the outpatients: those having consulted for this infection in the infectious disease department and who did not require hospital care until the end of the acute infectious episode. Data of interest were collected from inpatients hospitalized from March 23 to April 9, 2020 and from outpatients who consulted from February 28 to March 30, 2020.

This study is observational. The study has been approved by the ethics committee of Sorbonne University (2020 - CER-2020-13).

### Definitions and data collected

Confirmed COVID-19 was defined as a positive result on real-time reverse-transcriptase– polymerase-chain-reaction (RT-PCR) assay of nasal and pharyngeal swab specimens.

Smoking status was collected in all patients by specifically asking whether they were current smokers (and if so, to provide details on their smoking habits: daily or occasional smoking, number of daily cigarettes), former smokers, or not smokers ever. We used the same definition as in the French national annual survey of smoking habits (Santé Publique France Health Barometer)[14] Daily smokers were defined as individuals reporting daily smoking of cigarettes (manufactured or rolled) or other tobacco products (cigars, cigarillos, pipe, shisha). Occasional smokers were defined as individuals reporting infrequent, but not daily smoking. The group of former smokers included anyone having smoked in the past, occasionally or daily, and had abstained from smoking prior to COVID-19 onset. The term “never smoker” designated people who had never smoked.

In addition, for all outpatients and for all inpatients, we systematically asked former smokers since when they had quitted smoking, current smokers whether they quitted since the onset of COVID-19 symptoms, and if so, if they took nicotinic substitutes (including with e-cigarette), and former smokers whether they used nicotinic substitutes (including with e-cigarettes) at the time of COVID-19 onset of symptoms. We also asked non smoker oupatients whether they used nicotinic substitutes (including with e-cigarettes) at the time of COVID-19 onset of symptoms.

Finally, the following data were extracted from the medical charts: age, sex, healthcare workers or not, comorbidities, known to have potentially an impact on the prognosis of COVID-19, including diabetes, hypertension, obesity, immunodepression and COPD, and out- or inpatient status.

For inpatients, the following outcomes within one month following first day of clinical manifestation were also extracted:, still hospitalized in medical ward without any ICU stay, discharged without any ICU stay or if they occurred earlier: transfer to ICU and still alive at day 30, death (both in ICU or in medical wards).

### Smoking rates in the population of reference

The French general population was used as a reference to compute the Standardized Incidence Ratio (SIR). Rates of daily smokers in France have been reported for the year 2019 by sex and age class (of 10 years) from the French national Survey “Santé Publique France Health Barometer” [14], a cross sectional phone survey made yearly on a representative sample of 18-85 year-old people living in mainland France, with a on 2-level random sampling[14]. The 2019 survey involved a sample of 10,352 individuals. The completion of the survey took place from January 9 to June 29, 2019 and used the same definitions of daily smokers, occasional smokers, former smokers and never smokers as described above. Age and sex rates are reported only for current daily smokers (not for occasional current smokers, former smokers nor non-smokers) aged from 18 to 75 years, and the rate of current daily smokers in the 76-85 year-old people was reported globally and not by gender.

### Statistical analysis

A descriptive analysis has been made by group (inpatients - outpatients). Qualitative variables were described by numbers and percentages, and quantitative variables by median and interquartile range. Inpatients and outpatients were compared for age and sex with Wilcoxon test and Pearson Chi2 tests, and for comorbidities and smoking status by logistic regression adjusted on age and sex. The SIRs were used to compare daily smoker rates in the COVID-19 inpatients and outpatients, respectively, with those of daily smokers in a reference population, here the French general population in 2019. The estimated SIR and its 95% confidence interval is the ratio between the observed number of daily smokers among the COVID-19 patients and the number of daily smokers that would be expected in the study population, on the basis of age- and gender-specific current daily smokers rates in the general population. The main analysis involved all included patients, and those older than 75 years were considered in the 65-75 years age class for standardization (reference rates of daily smokers of 10.4% in men and 9.0 in women), which for our hypothesis is a conservative approach, because daily smoker rates decreases with age (4.8% of daily smokers within the 76-85 year-old people in France in 2019, but the rates are not available by sex). For 7 outpatients and 2 inpatients, we were unable to interview the patient on his smoking status. We did not include the latter patients in the main analysis because the missing smoking status was very likely to be at random (7 outpatients that could not be reached, and among the 2 inpatients, one due to the language barrier and the other due to severe cognitive impairment). We performed two sensitivity analyses, one excluding patients older than 75 years, the other considering the patients with missing smoking status as daily smokers.

We also estimated the SIR in healthcare workers and non healthcare workers in the outpatients (as healthcare workers were overrepresented, because they were tested at their workplace in case of symptoms).

## Results

### Demographic and Clinical Characteristics

A total of 340 inpatients and 139 outpatients were included. The demographic and clinical characteristics of the two groups are shown in TABLE 1. As shown in figure 1, age distribution differed between outpatients and inpatients, with outpatients being younger and inpatients older.

**TABLE 1:**
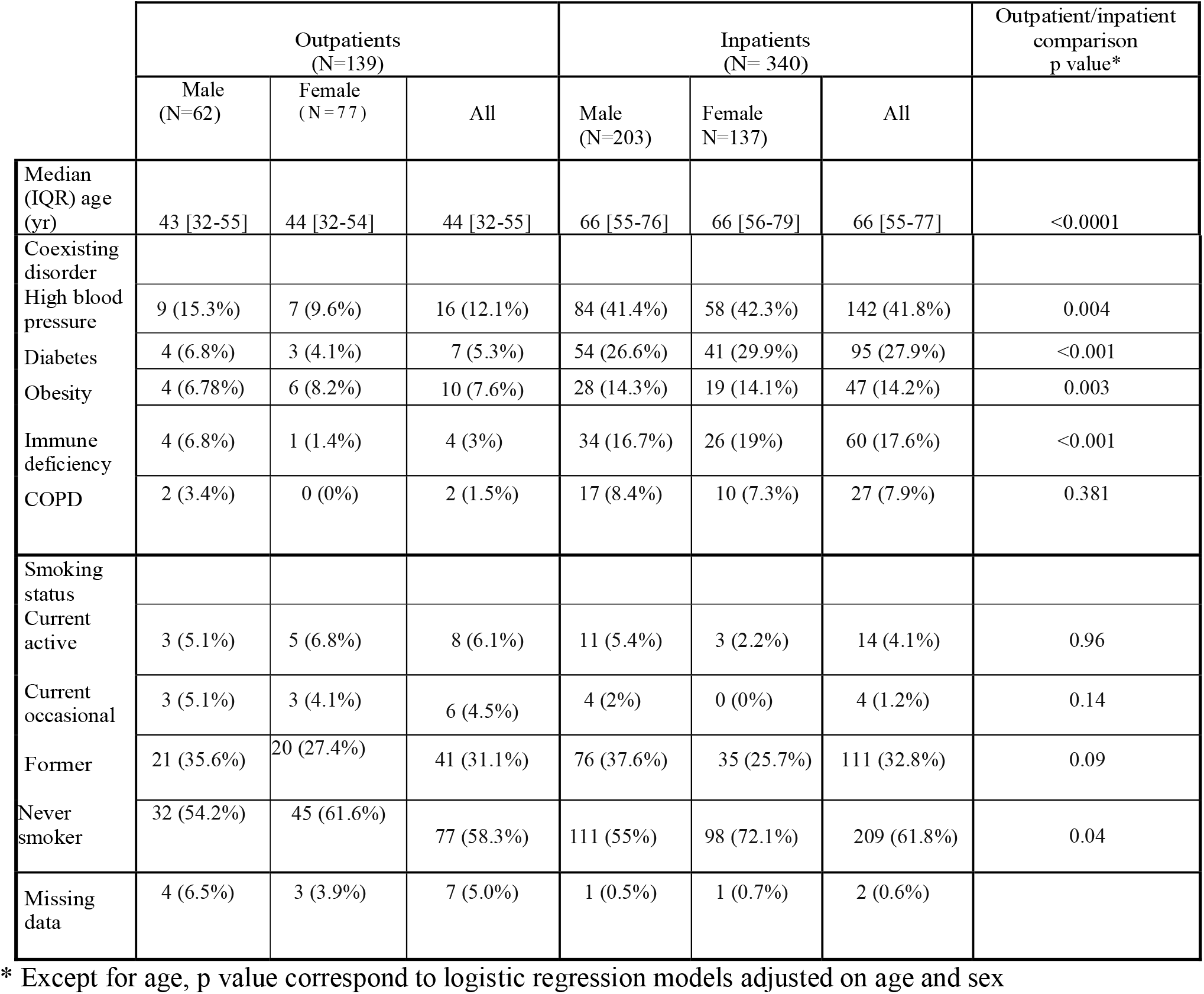
Clinical characteristics and smoking habits of COVID-19 patients

**Figure 1.**
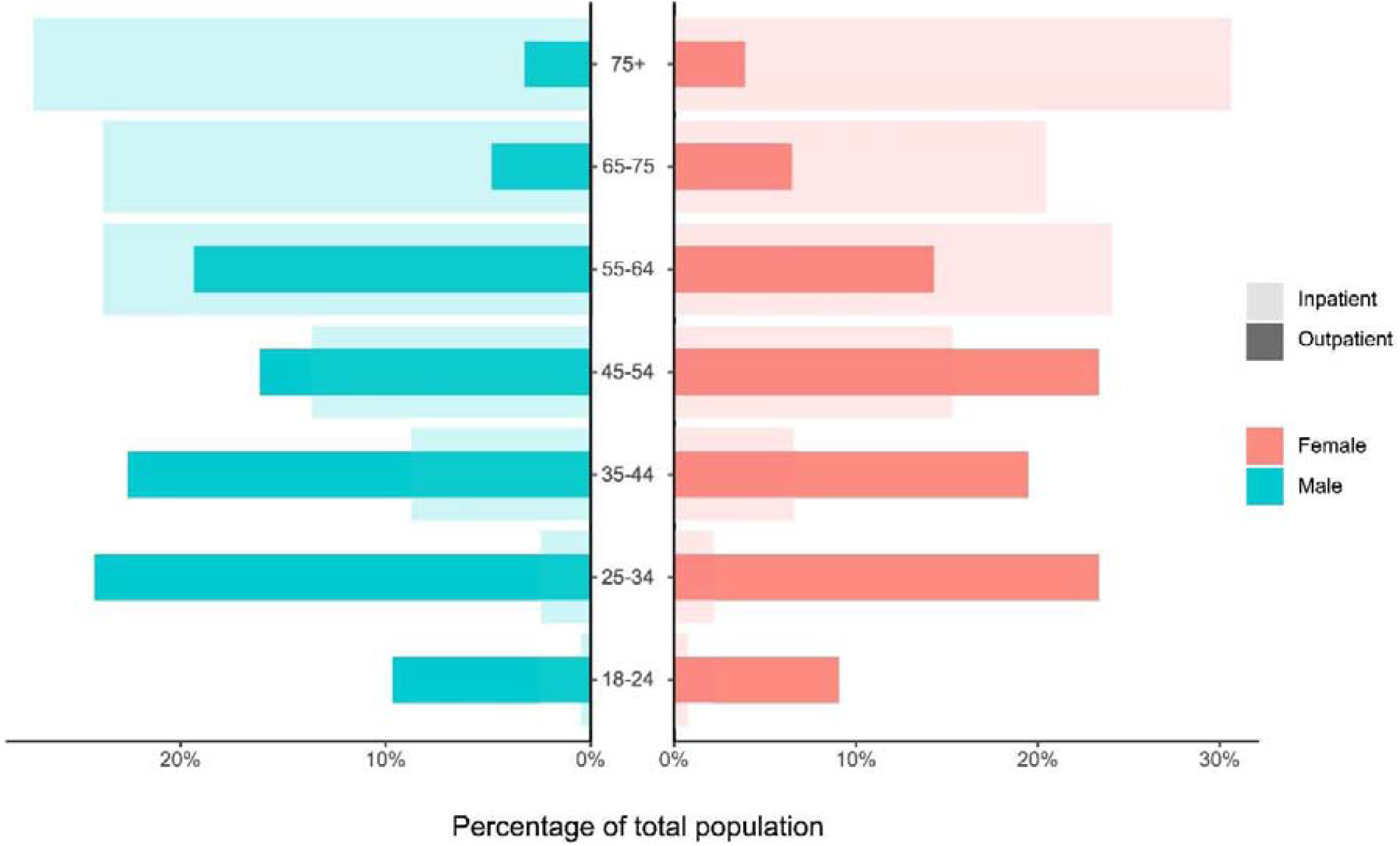
Age and sex distribution in COVID-19 inpatients and outpatients. Dark and light shaded histograms represent outpatients and inpatients with confirmed COVID-19 status, respectively

**Figure 2.**
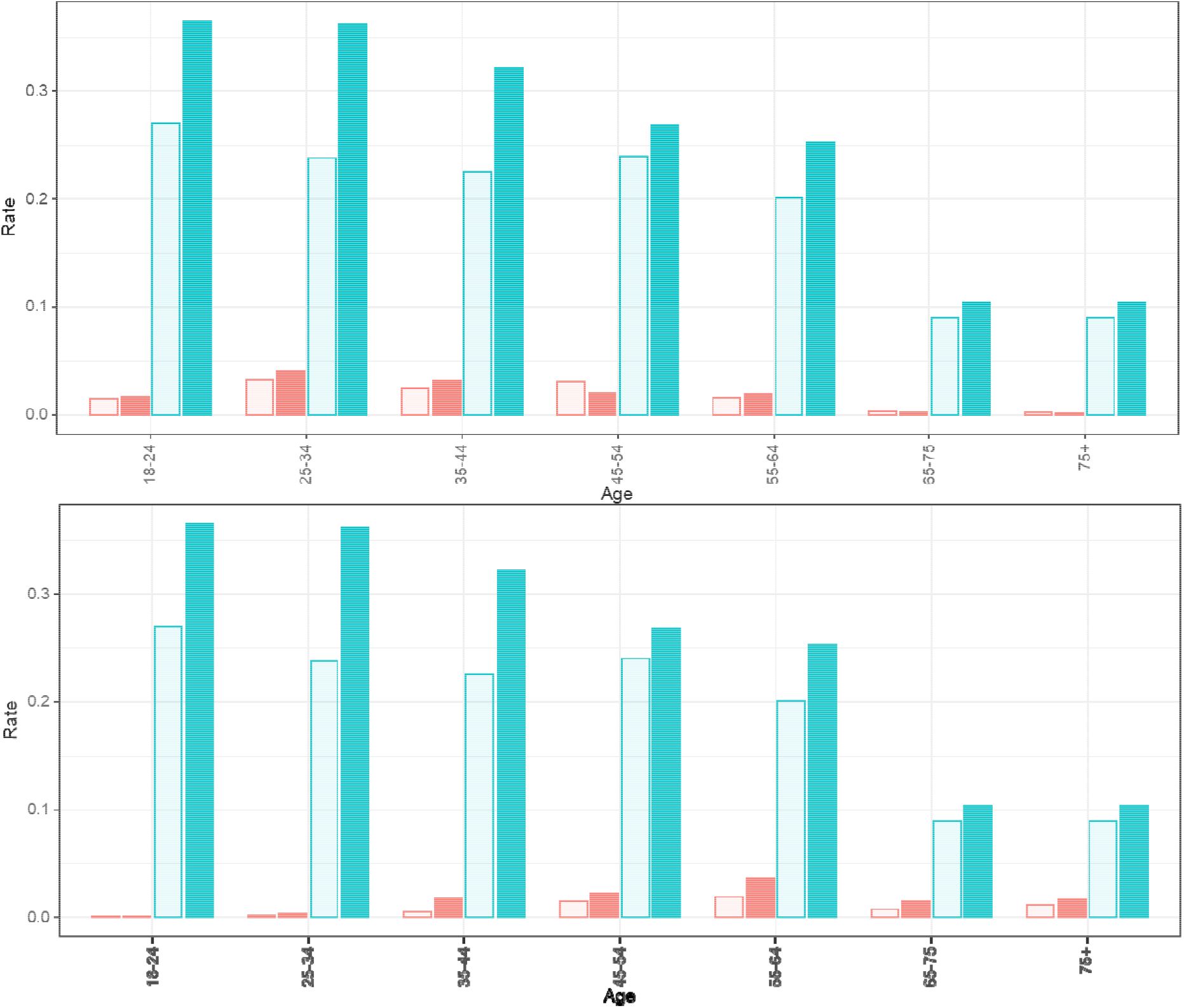
Age and sex expected rates of daily smokers in COVID-19 patients (A) For outpatients. (B) For inpatients Light shaded and dark histograms represent women and men daily smokers, respectively. In blue: expected incidence rate in each age and sex class; in red: expected incidence rate in each age and sex class.

The inpatient group was composed of 340 patients, median age 66 years: 203 men (59.7%, median age 66 years) and 137 women (40.3%, median age 66 years). The rate of daily smokers was 4.1% CI95% [2.3 – 6.9] (5.4% of men and 2.2% of women) corresponding to 14 patients. Among them, 4 smoked 5 cigarettes/day or less, 3 smoked 6 to 10 cigarettes/day, 1 smoked 15 cigarettes/day and 5 smoked 20 or more cigarettes/day (and the data was missing for 1). For former smoker inpatients (n=111, 32.8 %), time duration since quitting was available for all but 6 patients. Five (4.8%) patients had quitted 2 months, and 2 (1.9%) patients 6 months before the clinical onset of the disease and 98 (93.3 %) more than one year before disease onset. Two former smokers (1.9%) were using nicotine substitutes (one by e-cigarettes and one by patches) at the time of disease onset.

The outpatient group was composed of 139 patients, median age 44 years: 62 men (44.6 %, median age 43 years), and 77 women (55.4 %, median age 44 years). In all, 68 (51.5%) were healthcare workers. Smoking status was missing for 7 patients. The daily smokers rate was 6.1% CI95% [2.7 - 11.6] (5.1% of men and 6.8 % of women) corresponding to 8 outpatients. Among them, 3 smoked less than 5 cigarettes/day, 3 smoked 6 to 10 cigarettes/day, and 2 smoked 20 or more cigarettes/day. After COVID-19 onset, 2 have stopped smoking, and none have taken nicotinic substitutes. Occasional smokers were 6 (4.5%), 2 have stopped smoking since COVID-19 onset and none have taken nicotinic substitutes. Former smokers were 41 (31,1%; 21 men and 20 women). Among these, 2 (4.9%) had quitted three months before COVID-19 symptoms onset and 39 (95.1%) more than 1 year before; 2 (4.9%) were using nicotinic substitutes (1 by use of e-cigarette). Among the 77 non-smokers, none were using nicotinic substitute (data was missing for 7).

The comorbidities were more frequently observed in inpatients than in outpatients: hypertension (age and sex-adjusted OR : OR_adj_= 2.5; 95%CI(1.4-4.8); p=0.004), diabetes (OR_adj_=5.4; 95%CI(2.4-13.7); p<0.001), obesity (OR_adj_=3.7; 95%CI(1.7-8.9), p=0.002), immune deficiencies (OR_adj_==12.45; 95%CI(4.6-44.3); p<0.001) except for COPD (OR_adj_==2.0; 95%CI(0.5-13.3), p=0.38).

### Outcome of COVID-19 inpatients

The outcome of inpatients is described in TABLE 2 and was as follows: 211 discharges without any ICU stay (62.1%), 54 still hospitalized in medical ward without any ICU stay (15.9%) by one month after onset of clinical symptoms and 29 transfers to ICU (8.5%) and 46 deaths in ICU or medical wards (13.5%). Among the 14 daily smokers, 1 (7.1%) patient died and 1 (7.1 %) has been referred to intensive care unit by day 30 after clinical onset, while all occasional smokers were discharged. 23 former smokers (20.7 %) and 21 non smokers (10%) died while 11 former smokers (9.9%) and 17 non smokers (8.1%) have been transferred to ICU. Thus, active smokers represented 2.2% and 3.4% of the 45 dead patients and the 29 patients transferred to ICU respectively, whereas they represented 4.1% of the inpatients.

**TABLE 2:**
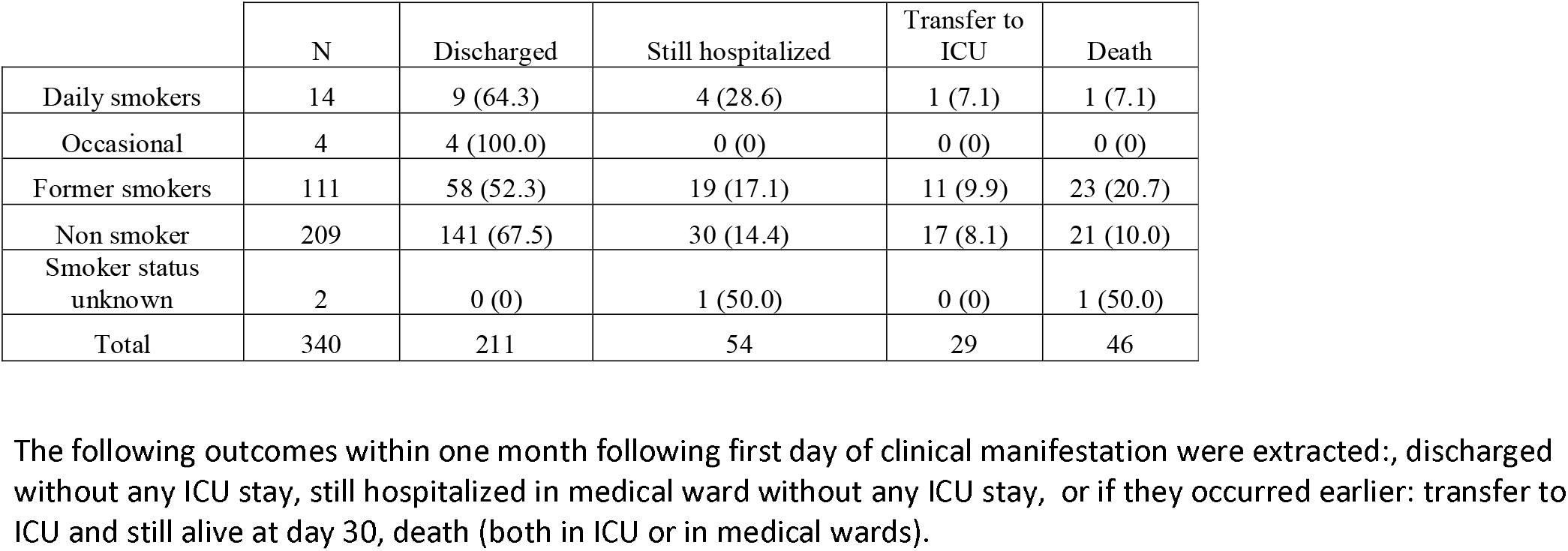
OUTCOME OF INPATIENTS

|  | N | Discharged | Still hospitalized | Transfer to ICU | Death |
| --- | --- | --- | --- | --- | --- |
| Daily smokers | 14 | 9 (64.3) | 4 (28.6) | 1 (7.1) | 1 (7.1) |
| Occasional | 4 | 4 (100.0) | 0 (0) | 0 (0) | 0 (0) |
| Former smokers | 111 | 58 (52.3) | 19 (17.1) | 11 (9.9) | 23 (20.7) |
| Non smoker | 209 | 141 (67.5) | 30 (14.4) | 17 (8.1) | 21 (10.0) |
| Smoker status unknown | 2 | 0 (0) | 1 (50.0) | 0 (0) | 1 (50.0) |
| Total | 340 | 211 | 54 | 29 | 46 |

The following outcomes within one month following first day of clinical manifestation were extracted: discharged without any ICU stay, still hospitalized in medical ward without any ICU stay, or if they occurred earlier: transfer to ICU and still alive at day 30, death (both in ICU or in medical wards).

### Comparison of the daily smoker rate with the French general population

The age and sex-SIR for daily smokers are shown in TABLE 3. In the main analysis, SIRs were 0.24 [0.12 - 0.48] and 0.24 [0.14 - 0.40] for outpatients and inpatients, respectively. The SIR in outpatients did not significantly differ from that in inpatients (p = 0.99). In the outpatients, the SIR was 0.17 [0.05-0.53] in the healthcare workers, and 0.32 [0.13-0.76] in the others. Sensitivity analyses yielded similar results.

**TABLE 3:**
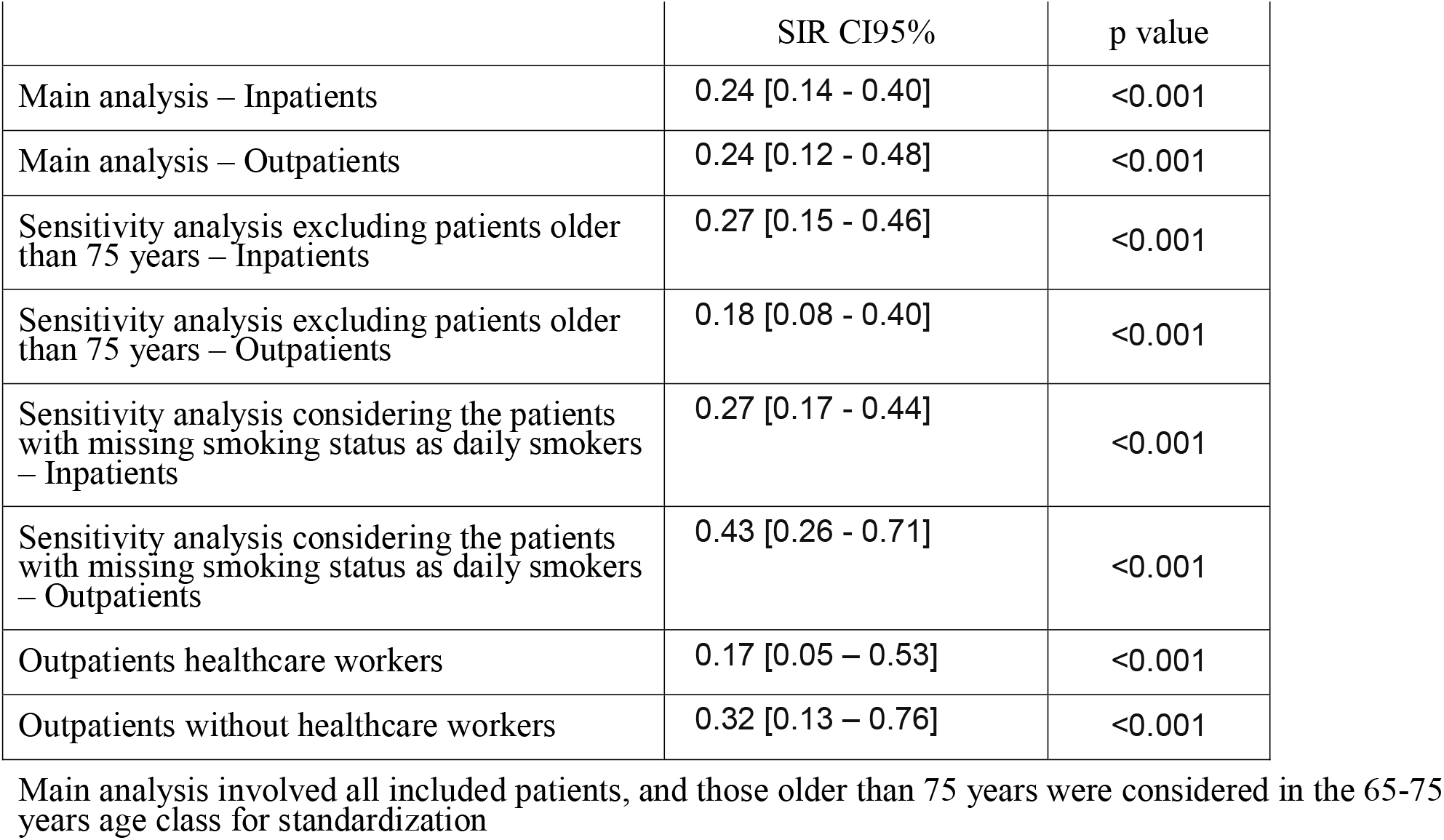
Standardized Incidence Ratios for daily smokers

|  | SIR CI95% | p value |
| --- | --- | --- |
| Main analysis – Inpatients | 0.24 [0.14 - 0.40] | <0.001 |
| Main analysis – Outpatients | 0.24 [0.12 - 0.48] | <0.001 |
| Sensitivity analysis excluding patients older than 75 years – Inpatients | 0.27 [0.15 - 0.46] | <0.001 |
| Sensitivity analysis excluding patients older than 75 years – Outpatients | 0.18 [0.08 - 0.40] | <0.001 |
| Sensitivity analysis considering the patients with missing smoking status as daily smokers – Inpatients | 0.27 [0.17 - 0.44] | <0.001 |
| Sensitivity analysis considering the patients with missing smoking status as daily smokers – Outpatients | 0.43 [0.26 - 0.71] | <0.001 |
| Outpatients healthcare workers | 0.17 [0.05 – 0.53] | <0.001 |
| Outpatients without healthcare workers | 0.32 [0.13 – 0.76] | <0.001 |

Main analysis involved all included patients, and those older than 75 years were considered in the 65-75 years age class for standardization

To note, the daily smoker rate in the 76-85 years inpatients and outpatients was 1.6% and 3.8%, respectively, lower than 4.8% observed in the French 76-85 years people in 2019 population.

## Discussion

This cross sectional study shows that the daily smokers rate is significantly lower in symptomatic COVID-19 patients than in the French general population, either for outpatients and inpatients. The SIRs of daily smokers in COVID-19 outpatients and inpatients were 0.24 [0.12 - 0.48] and 0.24 [0.14 - 0.40], respectively, which means a decrease of 76% as compared to the French population, accounting for age and sex distribution. This result suggests that daily smokers have a lower probability of developing symptomatic SARS-CoV-2 infection as compared to the general population. The SIRs did not differ between outpatients and inpatients, suggesting that the potential effect of smoking is towards symptomatic COVID-19, irrespective of the severity. In the rare daily smokers in the COVID-19 patients of our study, we did not observed any effect of the daily cigarette consumption. Actually some were heavy smokers and others not. To note, in 2019, the mean number of daily cigarettes by current smokers in the French general population was 12.5 cigarettes, or equivalent, with 13.5 cigarettes for men and 11.4 for women [14]. We also observed a very rare use of nicotinic substitutes in the former smokers (2/111 in the inpatients and 2/41 in the outpatients, one of each group with e-cigarette), and in none of the outpatients not-smokers, which is in line with the national survey indicating that e-cigarette use is still low in France (4.4% of daily users), and is not used by non smokers (1% of e-cigarette users).

Previous studies have reported low current smokers rate (or low smokers rate, with no distinction between current and former smokers) in COVID-19 patients, ranging from 1.4% to 12.6% in China (National Chinese current smokers rate was 52% in men, 2.5 % in women in 2015, and 27.3% of adults in 2018)[15], of 1.3% in the USA in the US Center of Disease Control report[11] and 5.1% in a report from New York city (US current smokers rate of 15.6% in men and 12% in women in 2018).[12, 16]

Our study investigates the smoking status of COVID-19 outpatients (non-severe cases) and inpatients (severe cases), separately. All previous studies but two reported smokers rates only in hospitalized patients[13], thus gave no information on whether low current smoker rates was related to severe (i.e. hospitalized) patients only, who have more frequently comorbidities and may have been strongly advised to quit smoking, or to any form of COVID-19. Smoking data from inpatients and outpatients were mixed in the Guan study[1]. The CDC, reported current smokers rates of 1.3% for the whole population of COVID-19 patients, 1% for outpatients, 2% for patients, not hospitalized in an ICU, and 1% in intensive care unit (ICU)-admitted patients[11], however, the level of missing smoking status was very high. In all previous studies, data were extracted from medical files.

Furthermore, in these previous studies, only crude smokers rates are reported, not compared to a control group or the general population except in two, where the current smokers rate in the general population is reported, with no statistical comparison and thus not accounting for the age and sex distribution of the COVID-19 patients.

Our findings are in line with those from Fontanet et al. 2020[17], who reported smoking habits in a cohort of pupils, their parents and siblings, as well as teachers and non-teaching staff of a high-school located in Oise (n = 661). Smokers had a lower risk of confirmed COVID-19 (as defined by antibodies detection) compared to non-smokers (7.2% vs 28.0; age-adjusted OR = 0.23; 95% CI = 0.09 – 0.59), and the association was also significant after adjustment on occupation.

Our study has many strengths. By contrast with previously reported studies, our study was specifically designed to assess smoking habits in the COVID-19 patients. Previous studies used smoking status as recorded in the medical files, which are subject to underreporting (usually not accounted as missing data) and biased reporting. In our study, patients were systematically interviewed about their smoking habits, and use of nicotinic substitutes.

The rate of missing data - one of the more frequent caveat of studies reported so far - was very low (1.9%). Additionally, to completely rule out the impact of missing data on the conclusion of our study, we did a sensitivity analysis, considering that patients with missing smoking status as daily smokers, which is conservative regarding the hypothesis of a protective effect of smoking. In this sensitivity analysis, the SIR remained significantly below 1 showing the robustness of our results. Furthermore, we used the same definitions as the French national annual survey of smoking categories (Santé Publique France Health Barometer)[14] that we used for reference to calculate the SIR. Finally, we investigated apart the association of daily smoking with COVID-19 separately in outpatients and inpatients, which provides relevant information in addition to previous studies.

Our study has also several limitations. First, the study was performed in early 2020 and the reference smoking rate in France were estimated from January to June 2019, as French smoking rates in 2020 are not available yet. However, it is very unlikely that a dramatic decrease in tobacco use may have occurred in France since mid 2019, which could explain our results. Actually, from 2017 to 2019, the daily smokers rate has decreased in France from 26.9% to 24.0%. The SIRs were estimated with the assumption that the studied population who lives in a limited area around a Parisian hospital has the same smoking habits as the general French population. Smoking rates are known to be lower in the Paris region (22.1% in 2017) than in other regions (26.9 % in France in 2017)[18], and this may have contributed to slightly overestimate the protective effect. Actually, smoking rates differ across socio-professional categories, and therefore may differ across geographic areas. It should also be noted that in the present study, healthcare workers were over-represented in the outpatient group, due to systematic testing at their work place when they become symptomatic, but not in the inpatient group. Health care worker represent an heterogeneous population with heterogeneous rates of smoking habits in France[19] and in other countries. In a systematic review and meta-analysis found an overall pooled prevalence of tobacco use in HCW of 21%, 31% in males and 17% in females[20] Additionally, even when estimating the SIR separately in healthcare and non healthcare oupatients, we still observed significantly lower daily smokers rates in the outpatients than in the general population. It is very unlikely that the very low SIRs that were estimated both for the out- and inpatient groups are the result of the study setting (we observed a 76% decrease in the COVID-19 population as compared to the French population, which is very substantial). Finally, because rates of occasional smokers, former smokers and of never smoker were not available by age and sex in the general population[14], we could not calculate SIR for these two smoker categories. However, on the hypothesis of the role of nicotine, only current smokers are concerned, and among them occasional smokers are scarce.

Second, because patients primarily hospitalized in ICU were not included in the present study, we could not conclude whether daily current smoking was associated or not with very severe forms of COVID-19. However, active smokers represented 2.2% and 3.4% of the 45 dead patients and the 29 patients transferred to ICU respectively, whereas they represented 4.1% of the inpatients, which is not in favor of a less favorable outcome in smokers. Furthermore as the rate of daily smokers was very low in both out- and inpatients, the study was not powered enough to assess whether smoking was associated with severity as defined by being hospitalized. However, it provides the information of a low smoking rate of daily smokers even in COVID-19 outpatients, which is of great interest in the understanding of the phenomenon. The association between daily smoking and COVID-19 severity still remains controversial[2]. A larger well-designed study including also ICU patients will certainly help to conclusively address this question. However, collecting accurately smoking status is difficult in ICU patients.

Third, smoking status was self-reported by the patients, which tend to underestimate daily smokers rate due to social desirability bias[21]. However, we used the same methodology as the Baromètre Santé survey that we used as reference. Furthermore, in the French healthcare system, access to care is not rationed based on any potential for positive outcome, or compliance with Public Health recommendations, thus there may be no particular incentive to underreport being a current smoker.

Another issue is that the low rate of smokers could be related to the association of COVID-19 with comorbidities leading to quit smoking. This limitation that could be discussed for inpatients does not however hold for outpatients who were mainly free of comorbidities.

Finally, in our study, smoking status was assessed only in symptomatic COVID-19 patients while a part of infected individuals are asymptomatic.[22] Thus we cannot conclude whether daily smoking is associated with SARS-CoV2 infection, or to symptomatic forms of this infection. The recent study by Fontanet[17], which highlights a decrease in the risk of COVID-19 of the same order of magnitude as us, provides an answer to this question because this study, based on serological results, takes into account both symptomatic and asymptomatic forms.

Because this is a cross-sectional study, we cannot conclude to the causality of the association. We cannot also identify which of the many compounds of tobacco exerts the protective effect of smoking on COVID-19. There are however, sufficient scientific data to suggest that smoking protection is likely to be mediated by nicotine. SARS-CoV2 is known to use the angiotensin converting enzyme 2 (ACE2) receptor for cell entry[23-25], and there is evidence that nicotine modulates ACE2 expression[26] which could in turn modulate the nicotinic acetyl choline receptor[27]. We hypothesize that SARS-CoV2 might alter the control of the nicotine receptor by acetylcholine. This hypothesis may also explain why previous studies have found an association between smoking and Covid-19 severity.[1, 3, 6] As hospitals generally impose smoking cessation and nicotine withdrawal at the time of hospitalization, tobacco (nicotine) cessation could lead to the release of nicotine receptors, that are increased in smokers, and to a “rebound effect” responsible for the worsening of disease observed in hospitalized smokers. However, this hypothesis needs further investigation, and the deleterious role of smoking in hospitalized patients with COVID19 cannot be ruled out to date.

In conclusion, our results suggest that active smokers may be protected against symptomatic COVID-19. This was evidenced for outpatients (who have less serious infections) as well as for hospitalized patients. The physiopathological process underlying this effect may involve nicotine through the nicotinic receptor (and not the smoke of cigarettes per se), a hypothesis that deserves further evidence. In light of the possible increased risk of severe form of COVID-19 among smokers once infected and of the long-term harmful consequences of smoking which is responsible for a very heavy public health burden with more than 78,000 deaths per year in France, our findings needs careful consideration and cannot be translating it into a clinical practice. Careful investigation of the potential protective effect of nicotine should be investigated both in *in vitro* and *in vivo* before any firm conclusion can be drawn.

## Data Availability

The data are available upon resquest to the corresponding author

